# Quantitative analysis of chest computed tomography of COVID-19 pneumonia using a software widely used in Japan

**DOI:** 10.1101/2023.06.22.23291669

**Authors:** Minako Suzuki, Yoshimi Fujii, Yurie Nishimura, Kazuma Yasui, Hidefumi Fujisawa

## Abstract

This study aimed to determine the optimal conditions to measure the percentage of area considered as pneumonia (pneumonia volume ratio, PVR) and the computed tomography (CT) score due to coronavirus disease 2019 (COVID-19) using the Ziostation2 image analysis software (Z2; Ziosoft, Tokyo, Japan), which is popular in Japan, and to evaluate its usefulness in assessing the clinical severity. We included 53 patients (41 men and 12 women, mean age: 61.3 years) diagnosed with COVID-19 using the polymerase chain reaction who had undergone chest CT and were hospitalized between January 2020 and January 2021. Based on the COVID-19 infection severity, the patients were classified as mild (n=38) or severe (n=15). For 10 randomly selected samples, the PVR and CT scores by Z2 under different conditions and the visual simple PVR and CT scores were compared, and the conditions with the highest statistical agreement were determined. The usefulness of the clinical severity assessment based on PVR and CT scores using Z2 under the determined conditions was statistically evaluated. The best agreement with the visual measurement was achieved by the Z2 measurement condition of ≥ –600 HU. The areas under the receiver operating characteristic curves, the Youden index, and the sensitivity, specificity, and p-values of PVR and CT scores by Z2 were as follows: PVR; 0.881, 18.69, 66.7, 94.7, and <0.001, CT score; 0.77, 7.5, 40, 74, and 0.002, respectively. We determined the optimal condition for assessing the PVR of COVID-19 pneumonia using Z2 and demonstrated that the AUC of PVR was higher than that of the CT score in the assessment of clinical severity. The introduction of new technologies is time-consuming and expensive; our method has high clinical utility and can be promptly used in any facility where Z2 has been introduced.

## Introduction

The coronavirus disease 2019 (COVID-19) pandemic caused by the novel coronavirus severe acute respiratory syndrome coronavirus 2 (SARS-CoV-2) was first identified in Wuhan, China, and reported in December 2019 [1]. The pandemic prevails with an increasing number of infections and deaths. In Japan, the first COVID-19 case was reported in January 2020 [2], and by April 2023, over 33 million people had been infected, and more than 74000 people had died [3]. After the Omicron strain of SARS-CoV-2 became prevalent, the number of severe cases complicated by pneumonia decreased, and the vaccination had spread socially. In May 2023, the legal classification was changed, and it was decided that a COVID-19 infection would be treated on the same level as an influenza virus infection [3].

This study was conducted with the aim of determining how the radiology department of a city hospital in Japan could use existing image analysis software to contribute to clinical practice at a time when the pre-Delta strain COVID-19 virus was predominant.

During the study period, COVID-19 infection had a high complication rate with pneumonia, especially in older adults [4, 5], with a high rate of aggravation and mortality, and it became necessary to distribute limited medical resources. The discrimination between mild and severe cases at the emergency department was an important and burdensome task. Typically, the severity was determined by symptoms, age, complications, blood tests, and computed tomography (CT) findings. The CT findings were generally evaluated visually, and the CT scores based on visual evaluation were not accurate or objective and took time and effort on the part of the evaluator. There are many reports on the CT severity assessment of COVID-19-associated pneumonia using an imaging software. The measurement methods and evaluation conditions differ for each individual tool, and few of them have been widely adopted in clinical settings.

The Ziostation2 image analysis software (Z2; Ziosoft, Tokyo, Japan) had been introduced in approximately 300 facilities in Japan, which was designed to quantify pulmonary emphysema in patients with chronic obstructive pulmonary disease. When a region above a certain concentration is recognized as a pneumonia region, the pneumonia volume ratio (PVR) can be measured by changing the threshold setting of the CT value (Fig 1a, b). To the best of our knowledge, there are no reports of COVID-19 pneumonia assessment by Z2.

**Fig 1.**
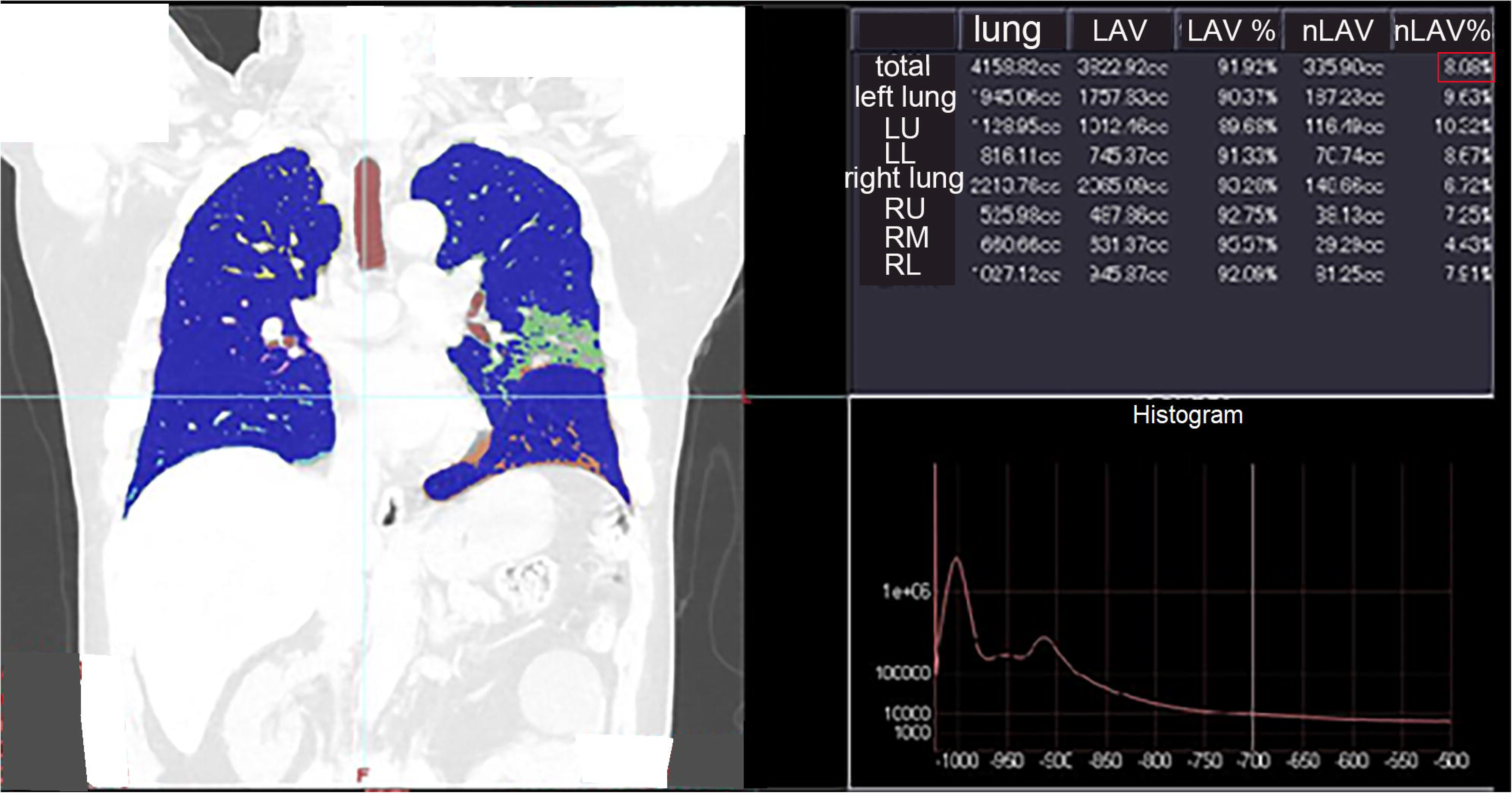
Images displayed on the console of the Z2. Z2 monitor screen. The PVR above a certain concentration is displayed in the upper right corner (red square). LAV, low attenuation volume; LL, left lower lobe; LU, left upper lobe; nLAV, not LAV (lung volume other than LAV); PVR, pneumonia volume ratio; RL, right lower lobe; RM, right middle lobe; RU, right upper lobe; Z2, Ziostation2.

Since Z2 has not been set to evaluate pneumonia, it is necessary to determine the threshold in Hounsfield units (HUs) for it. Therefore, it was decided to set the threshold at the concentration that most closely matched the visual evaluation.

In this study, we determined the appropriate conditions for the evaluation of COVID-19 pneumonia by Z2 through comparison with visual evaluation results and examined the usefulness of the clinical severity assessment of Z2.

## Materials and Methods

### Study population

This study adhered to the principles of the Declaration of Helsinki and was approved by the Ethical Review Committee of Fujisawa City Hospital (approval number: F2021022). The study was conducted retrospectively using imaging data and electronic medical records. An informed consent was provided by all patients in an opt-out manner on the website.

We evaluated patients diagnosed with COVID-19 using a polymerase chain reaction test who required a chest CT scan at our hospital and inpatient hospital care between January 2020 and January 2021. The patients who had an initial CT scan at another hospital or those who were initially treated at another hospital and subsequently transferred to our hospital, and cases without pneumonia findings on chest CT were excluded. Ten samples were randomly selected from patients under 65 years of age and with an uncomplicated condition.

The clinical severity of COVID-19 was classified as mild (SpO_2_ > 93%) or severe (SpO_2_ ≤ 93%, intubation, and intensive care unit management) based on the symptoms at the time of hospitalization, according to the guidelines of the Ministry of Health and Welfare [4]. The clinical severity, symptoms, comorbidities, blood test values, and clinical course were retrieved from the electronic medical records.

### CT protocol

The chest CT scans were obtained using 64-multidetector CT scanners (SOMATOM Definition AS 64; Siemens Healthineers, Erlangen, Germany). The CT parameters used at our hospital were as follows: 120 kVp, 160-316 mA current intelligent control (auto mA), and 5 mm slice thickness reconstruction. All CT examinations were performed without the use of intravenous contrast agents. The EV Report picture archiving and communication system (PACS) (PSP Corporation, Tokyo, Japan) was used to evaluate the CT findings.

### CT image analysis

Two radiologists evaluated the CT findings of pneumonia in all patients (Y.N. and M.S.) in consultation for the presence or absence of ground-glass opacity (GGO) (–/+), crazy-paving finding (–/+), consolidation (none/mild/moderate/severe), and emphysema (–/+).

For the 10 selected participants, visual evaluation of the PVR was performed independently by two radiologists (Y.F. and M.S.) using the free-form curve drawing tool of the PACS by adding up the area of the lungs and the pneumonia area freehand at 1.5-cm intervals in the coronal chest CT images (Fig 2). In the same participants, the two radiologists independently scored the percentage of pneumonia area in each lobe using visual measurements (0: 0%, 1: 25%, 2: 25–50%, 3: 50–75%, and 4: 75–100%).

**Fig 2.**
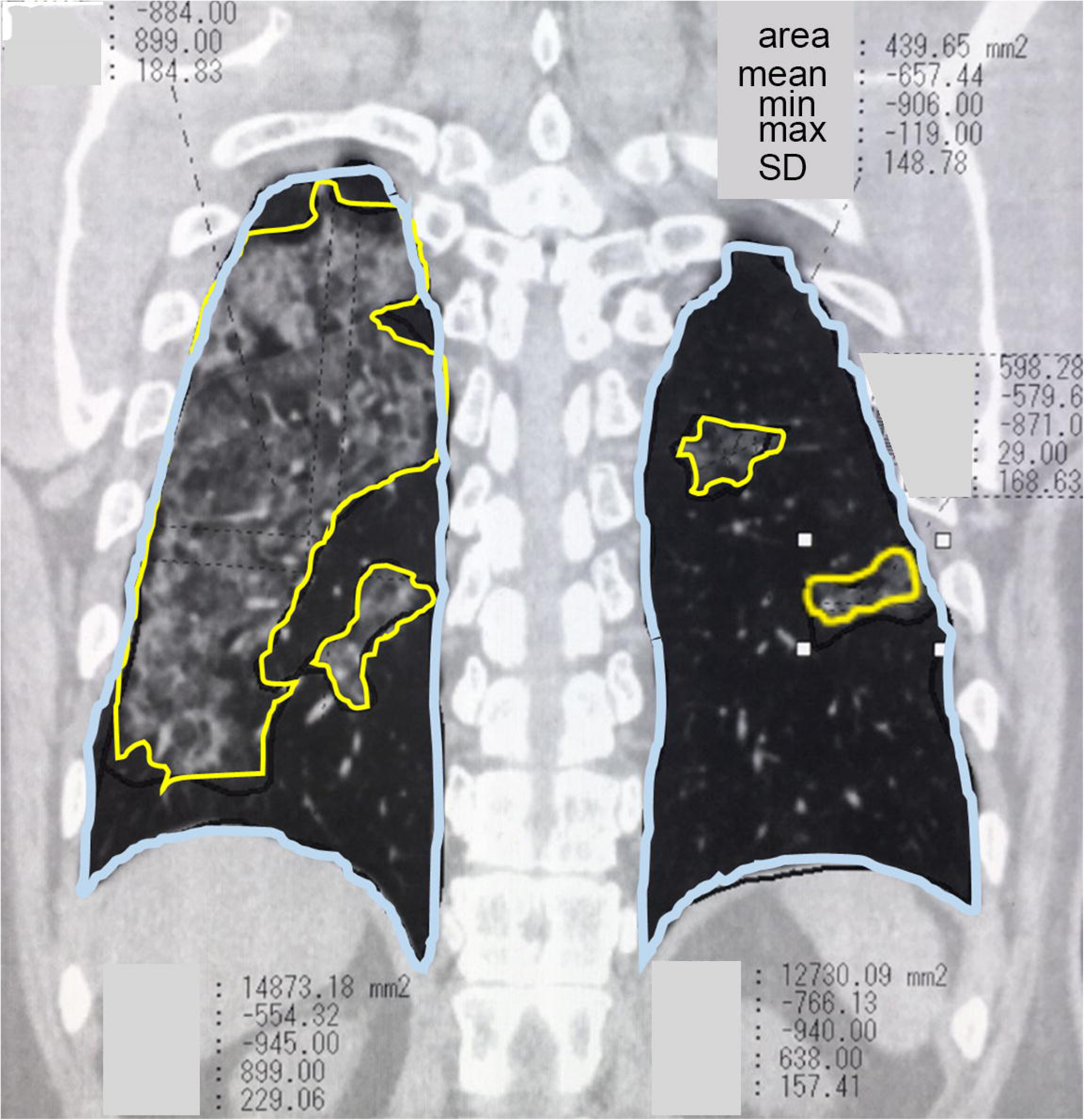
Visual measurement of PVR. Two radiologists independently selected the entire lung field and pneumonia area every 1.5 cm on the coronal view using a drawing tool on the PACS (PSP Corporation, Tokyo, Japan), and added these up to measure the PVR. The blue line indicates the entire lung field (mm^2^), and the yellow line indicates the pneumonia area (mm^2^). The minimum and maximum in the figure represent CT values in the region. PACS, report picture archiving and communication system; PVR, pneumonia volume ratio; min, minimum; max, maximum; SD, standard deviation.

Z2 provided the quantification of the emphysema, healthy lung parenchyma, GGO, and consolidation based on a HU. Z2 can divide segments and calculate total volumes for both the right and left lungs. In the measurement of PVR and CT scores in the 10 selected participants using Z2, the lung fields above a particular concentration were set as pneumonia areas and measured at ≥ –500 HU, ≥ –550 HU, ≥ –600 HU, ≥ –650 HU, and ≥ –700 HU. Z2 may not recognize the subpleural consolidation area as a lung field, and the total lung volume may be underestimated (Fig 3); therefore, radiologist A (M.S.) made the appropriate corrections manually.

**Fig 3.**
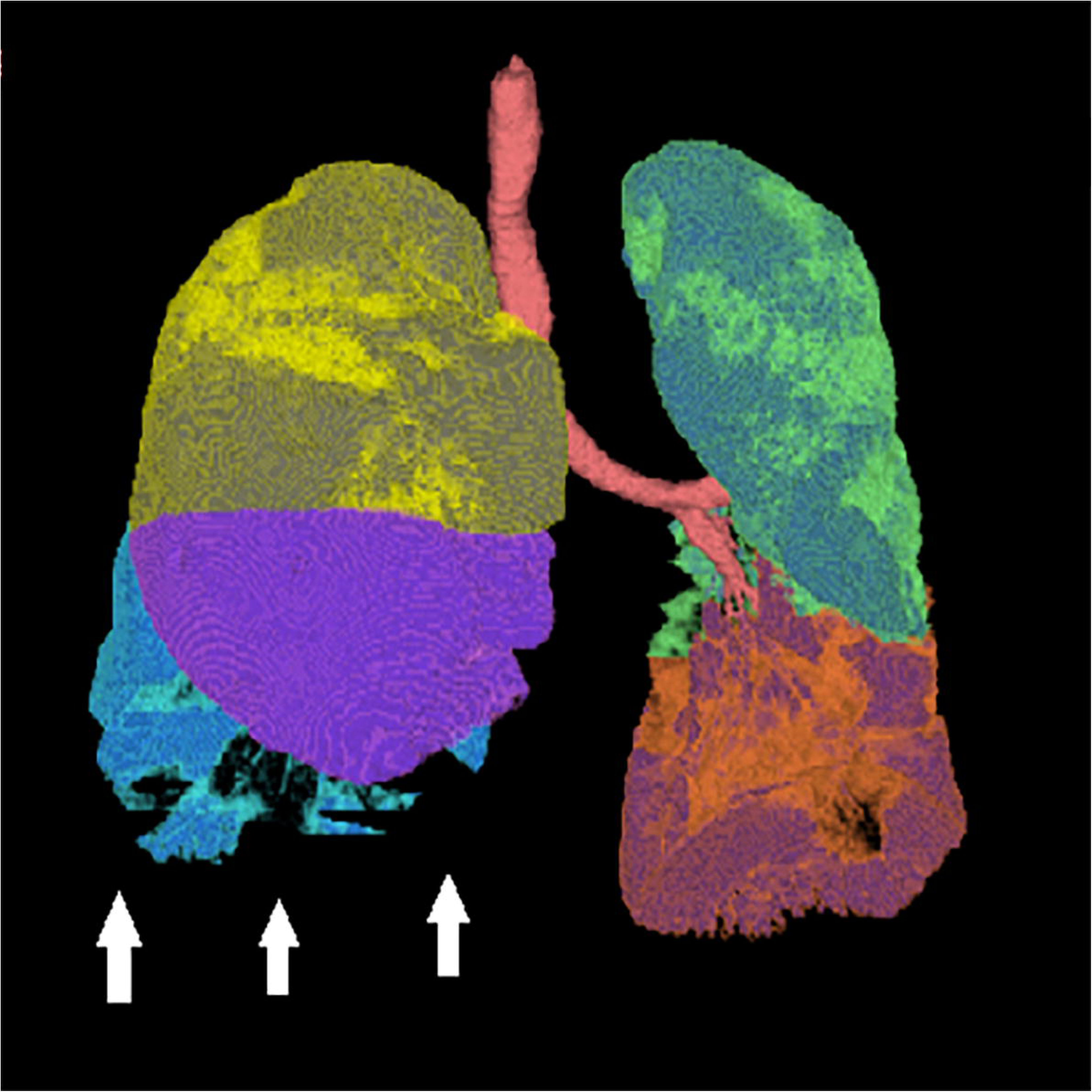
Dorsal subpleural consolidations are not recognized as part of the lung and require manual correction. The white arrows indicate the areas that needed to be manually corrected.

### Statistical analysis

The presence of significant differences in participant background (age, sex, number of days from disease onset to CT evaluation, and laboratory test results) between the mild and severe groups was evaluated using the t-test and chi-square test. The accuracy between the gross measurements of PVR and CT scores by two independent radiologists and the measurements by Z2 were evaluated using the Spearman’s rank correlation coefficient. The influence of possible confounding factors of participant background (age, sex, number of days from disease onset to CT evaluation, and presence of comorbidities) on the severity classification of PVR by Z2 was evaluated using the bivariable logistic regression. The usefulness of PVR and CT scores by Z2 under the determined conditions, primary laboratory tests, and CT findings in the clinical severity assessment was determined by the receiver operating characteristic (ROC) curves, Youden index, sensitivity, specificity, and p-values. All statistical analyses were performed using the SPSS software (version 27; IBM, Armonk, NY, USA).

## Results

The number of patients diagnosed with COVID-19 using a polymerase chain reaction test who required a chest CT scan at our hospital and inpatient hospital care between January 2020 and January 2021 were 91. Of these, three patients who received initial treatment at another hospital and 28 patients who had no findings of pneumonia on chest CT were excluded. Two cases were excluded from the study because the thin slice data necessary for Z2 measurement were not saved, and five cases could not be measured by Z2 for unknown reasons.

In total, 53 participants (41 men and 12 women, with a median age of 61.3 years; 38 in the mild group and 15 in the severe group) were included. Table 1 shows the participants’ demographics (age, sex, and presence of comorbidities), laboratory findings, and CT findings. Fifty-two participants presented with COVID-19 symptoms; however, there was no significant difference in the severity of the symptoms between the mild and severe disease groups. Significant differences in the number of days from disease onset to CT evaluation and the presence of comorbidities were found between the two groups. In addition, laboratory results revealed that C-reactive protein (CRP) and lactate dehydrogenase (LDH) levels differed significantly between the two groups. The CT findings showed a significant difference in consolidation between the two groups.

**Table 1.**
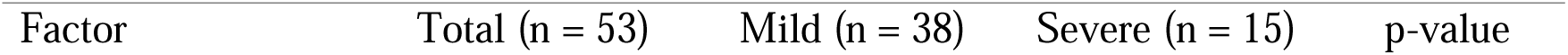
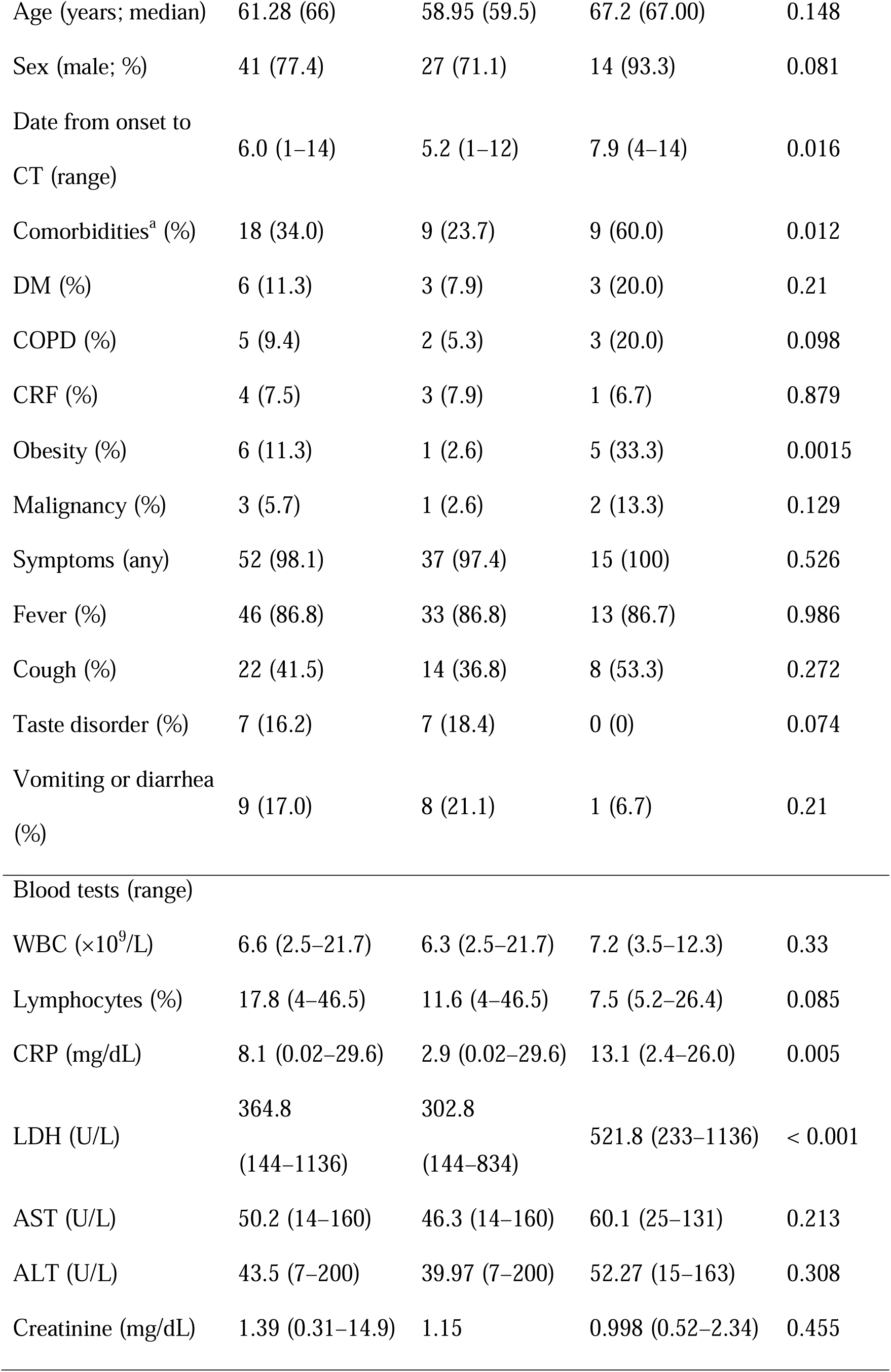
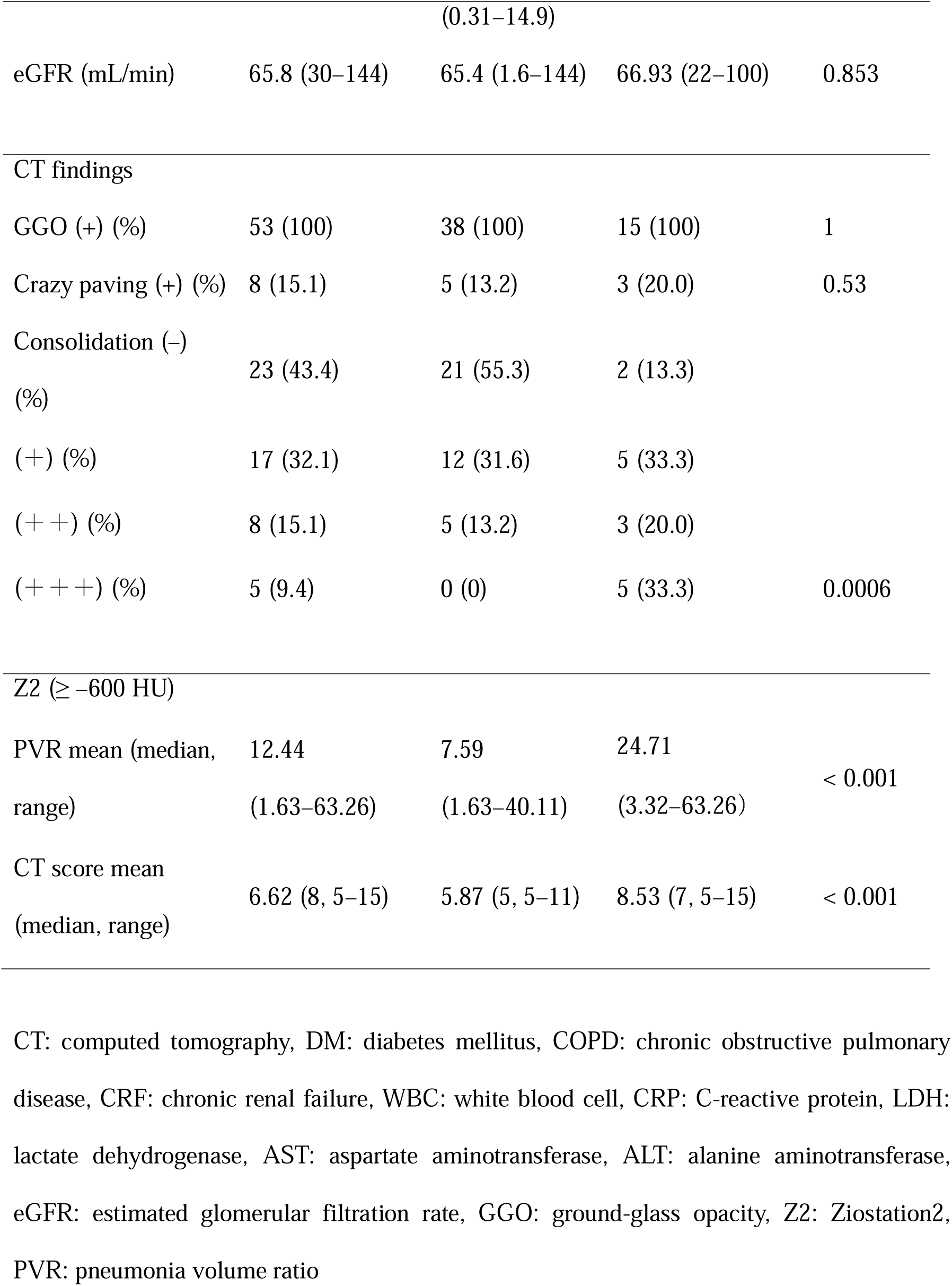
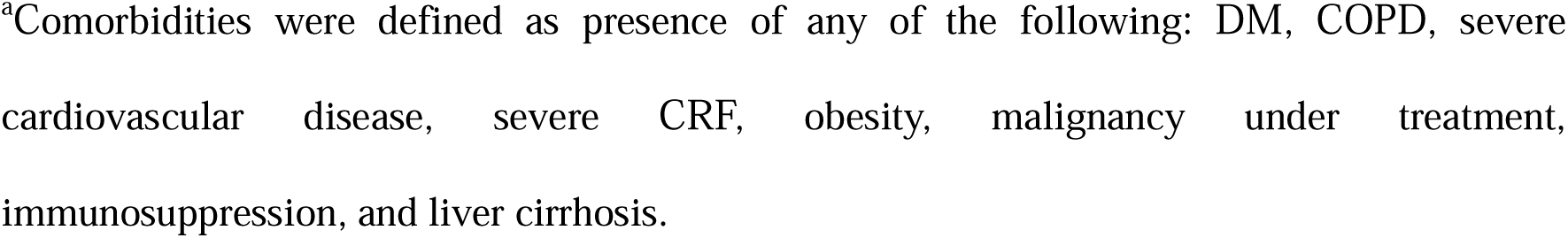
Patient background, blood test, and CT findings

Table 2 shows the results of the Spearman’s correlation between Z2 (under each condition; PVR: ≥ –500 HU, ≥ –550 HU, ≥ –600 HU, ≥ –650 HU, and ≥ –700 HU, CT score: ≥ –500 HU, and ≥ –600 HU) and the two radiologists for PVR and the CT scores in the 10 participants without comorbidities, respectively. While the accuracy between the two radiologists and Z2 for PVR was equally high at ≥ –500 HU to ≥ –600 HU, the accuracy for CT scores was higher at ≥ –600 HU than at ≥ –500 HU. Based on these results, the Z2 measurement condition for COVID-19 pneumonia that achieved the best accuracy with the gross measurement was determined to be ≥ –600 HU.

**Table 2.** Results of the Spearman’s test of PVR and CT score by two radiologists and Ziostation2 of five/two conditions in the 10 selected patients

| | | Reader | $\geq -500$ | $\geq -550$ | $\geq -600$ | $\geq -650$ | $\geq -700$ |
| --- | --- | --- | --- | --- | --- | --- | --- |
|  |  | B | HU | HU | HU | HU | HU |
| Reader A | PVR | 0.879 | 0.976 | 0.976 | 0.976 | 0.964 | 0.818 |
|  | CT score | 0.976 | 0.639 |  | 0.651 |  |  |
| Reader B | PVR |  | 0.842 | 0.842 | 0.842 | 0.83 | 0.661 |
|  | CT score |  | 0.584 |  | 0.696 |  |  |

PVR: pneumonia volume ratio, CT: computed tomography,

Reader A: M.S., Reader B: Y.F.

Figs 4 and 5 show the ROC curves and boxplots corresponding to the classification of disease severity by PVR and CT score using Z2 (≥ –600HU), CRP, and LDH. The areas under the curve (AUCs) were 0.881, 0.77, 0.788, and 0.842, respectively.

**Fig 4.**
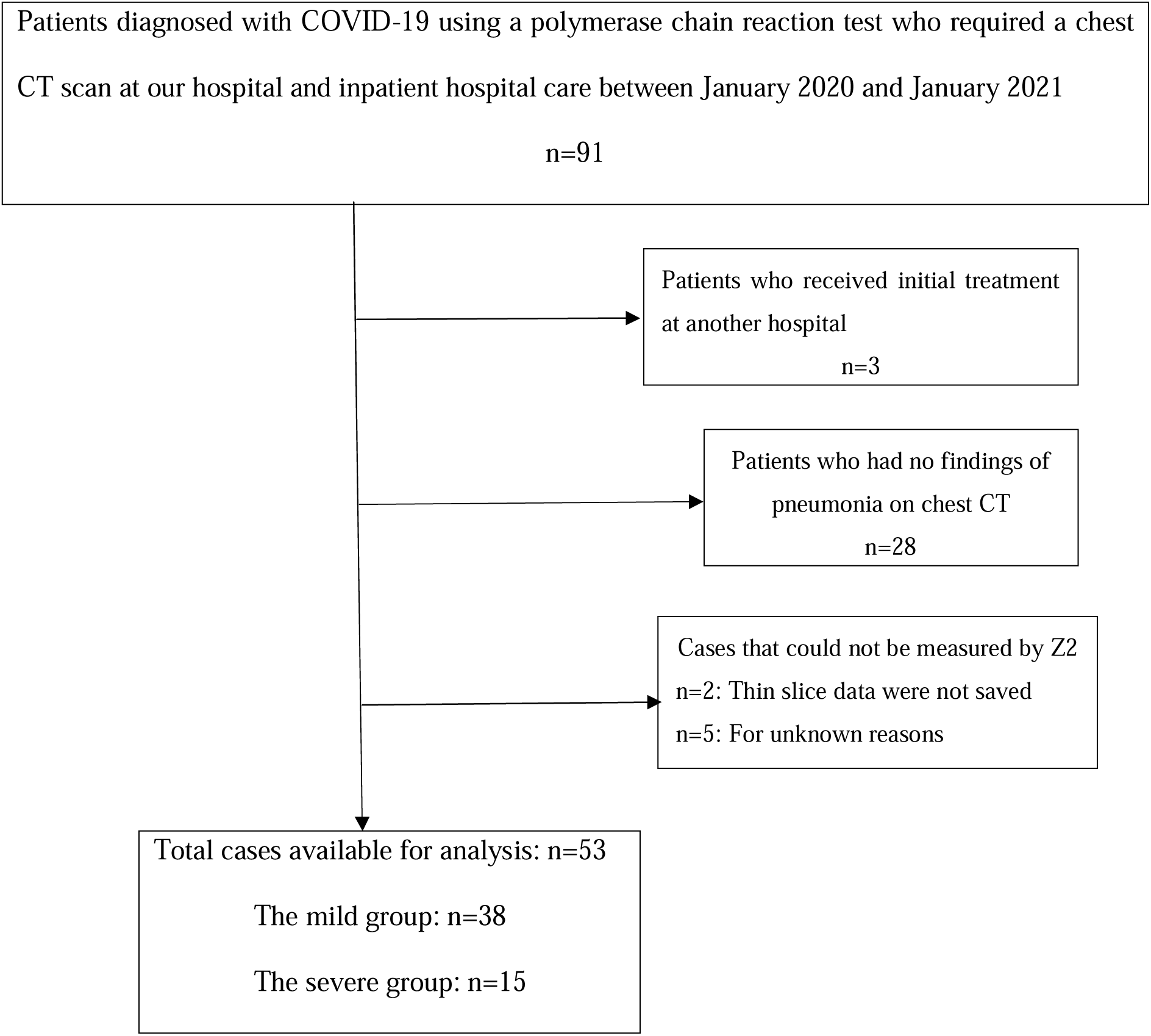
The flow chart shows the process of determining the number of study cases to 53.

**Fig 5.**
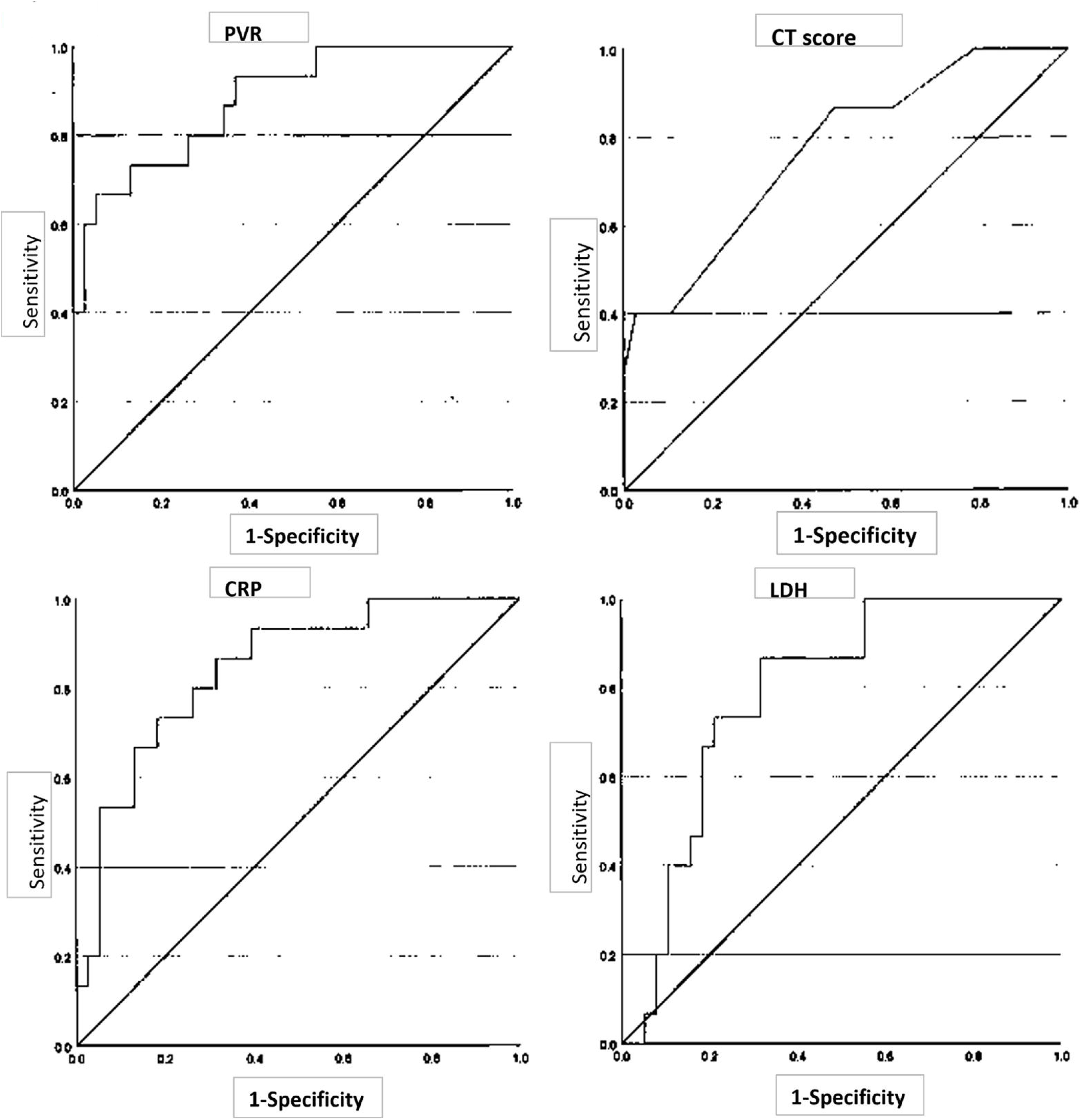
ROC curves for PVR, CT score, CRP, and LDH. ROC curve for **a**. PVR using Z2 (≥ –600 HU) and **b**. CT scores using Z2 (≥ –600 HU), **c**. CRP, and **d**. LDH. ROC, receiver operating characteristic; PVR, pneumonia volume ratio; Z2, Ziostation2; CT, computed tomography; CRP, C-reactive protein; LDH, lactate dehydrogenase; SD, standard deviation

**Fig 6.**
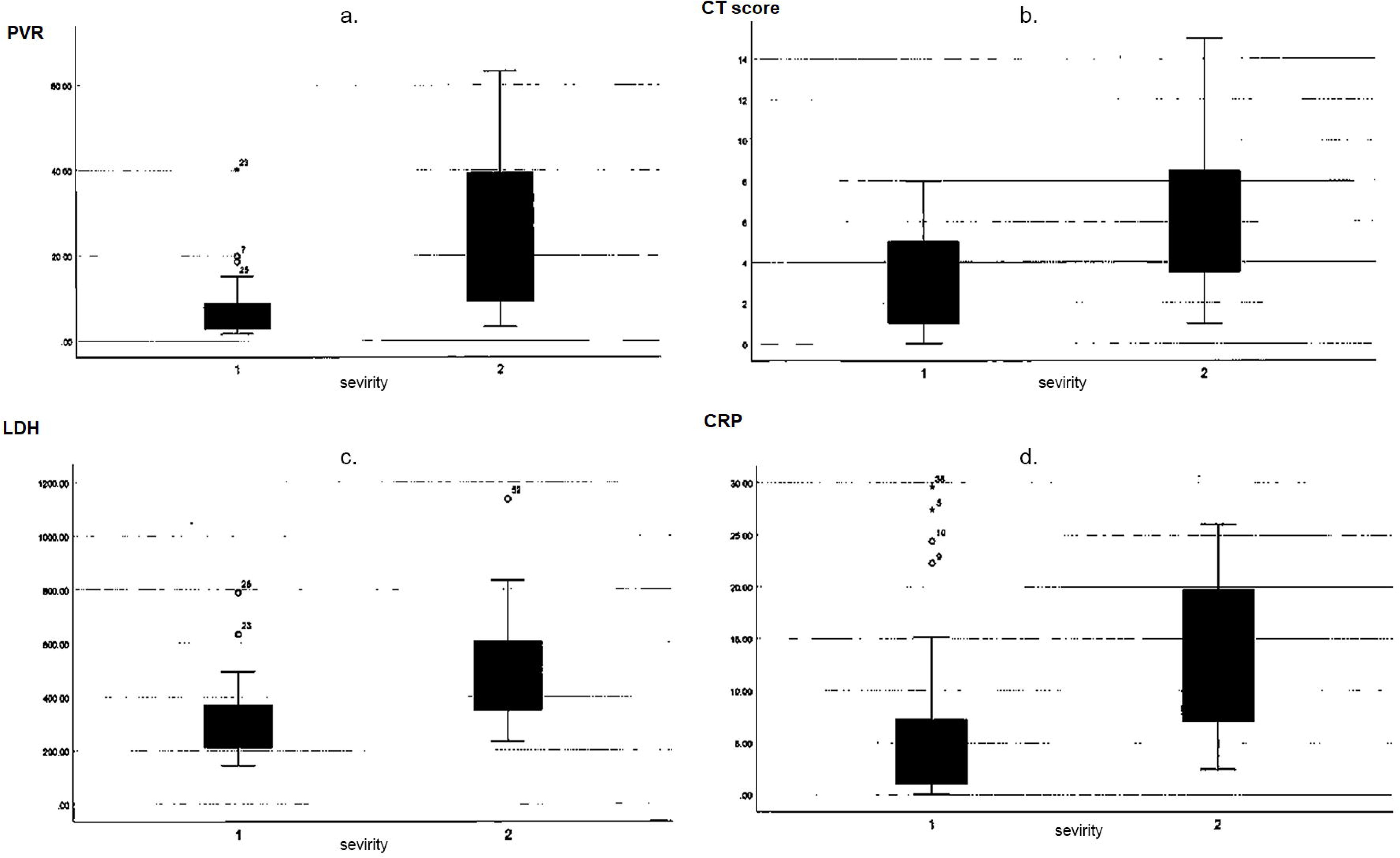
Boxplots of PVR, CT score, and CRP. Boxplots for **a**. PVR using Z2 (≥ –600 HU), **b**. CT scores using Z2 (≥ –600 HU), **c**. CRP, and **d**. LDH. 1: mild group, 2: severe group. Error bars indicate outliers. PVR, pneumonia volume ratio; CT, computed tomography; Z2, Ziostation2; CRP, C-reactive protein; LDH, lactate dehydrogenase

The Youden index values for PVR and CT scores at ≥ –600 HU by Z2, CRP, and LDH were 18.69, 7.5, 5.26, and 306.5, respectively. The sensitivities for PVR and CT scores at ≥ –600 HU by Z2 were 66.7% and 40%, respectively. The specificities for PVR and CT scores at ≥ –600 HU by Z2 were 94.7% and 74%, respectively. The p-value for PVR at ≥ –600 HU by Z2 was p < 0.001, and that for CT scores at ≥ – 600 HU by Z2 was p = 0.002 (Table 3). The bivariable logistic regression of PVR (≥–600 HU) according to age, sex, date from onset to CT, and comorbidities showed no significant effects, except for comorbidities (Table 4). The sensitivity and specificity were 66.7% and 89.5% when the PVR threshold was 18, and 60% and 97.4% when the PVR threshold was 20, respectively.

**Table 3.** Cut-off values for pneumonia volume ratio and blood test to differentiate mild.

|  | Youden<br>index | AUC | Sensitivity | Specificity | Lower<br>95%<br>CI | Upper<br>95% CI | p-value |
| --- | --- | --- | --- | --- | --- | --- | --- |
| PVR ($\geq -600$ HU) | 18.69 | 0.881 | 66.7 | 94.7 | 0.781 | 0.981 | < 0.001 |
| CT score ($\geq -600$ HU) | 7.5 | 0.77 | 40 | 74 | 0.629 | 0.911 | 0.002 |
| CRP | 5.26 | 0.788 | 86.7 | 68.4 | 0.664 | 0.912 | < 0.001 |
| LDH | 306.5 | 0.842 | 86.7 | 68.4 | 0.729 | 0.956 | < 0.001 |

AUC: area under the curve, PVR: pneumonia volume ratio, CT: computed tomography, CRP: C-reactive protein, LDH: lactate dehydrogenase, CI: confidence interval

**Table 4.**
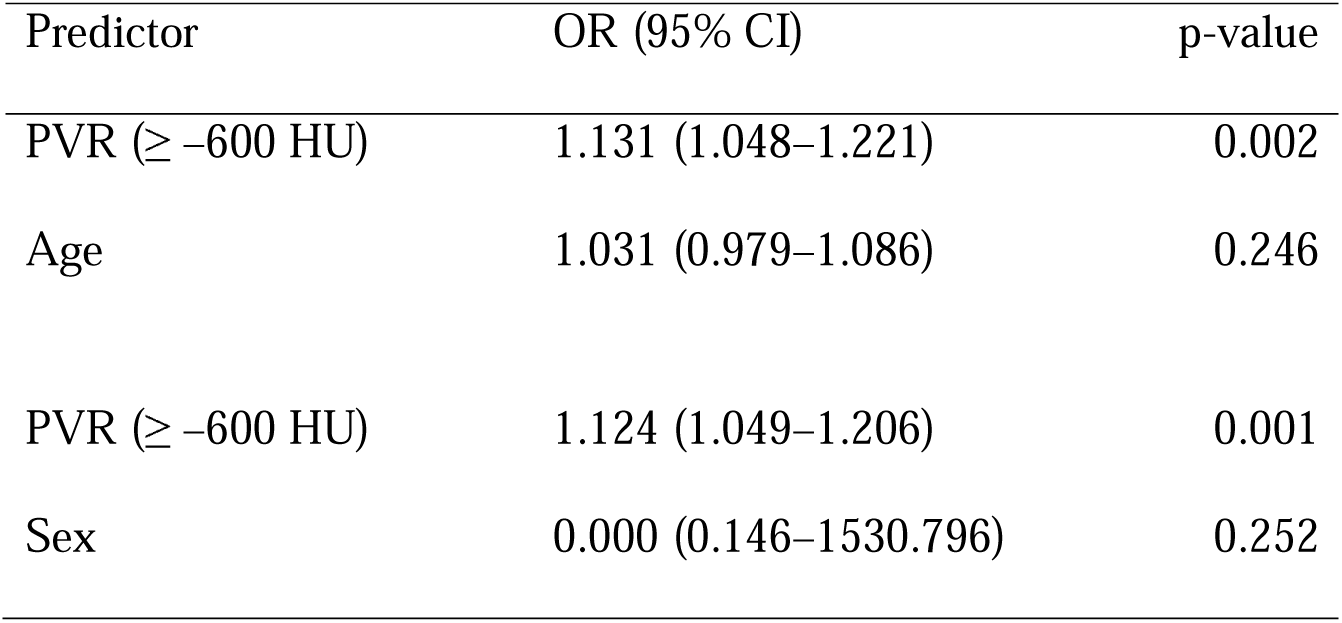
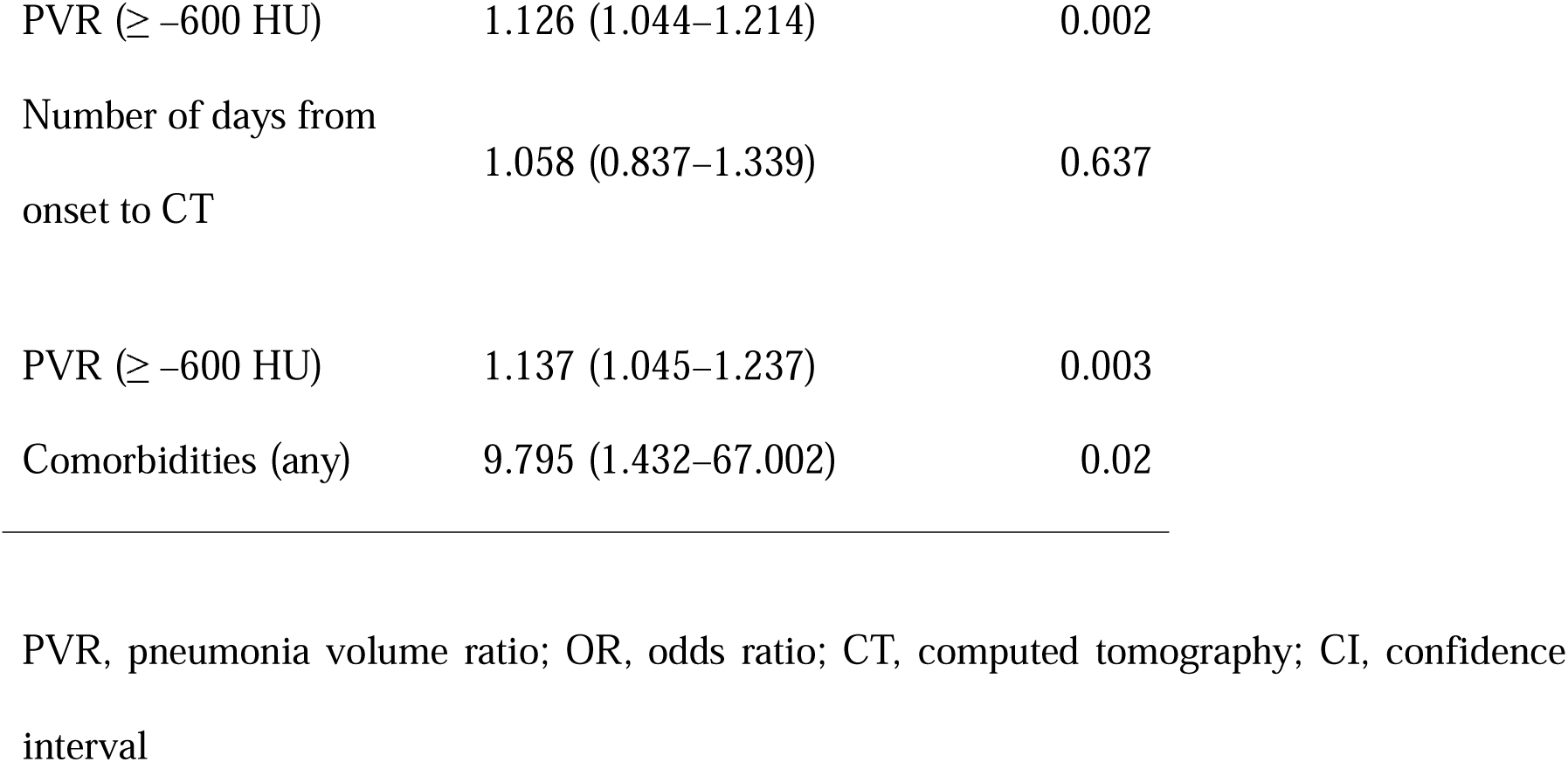
Bivariable logistic regression of PVR (≥–600 HU) according to age, sex, number of days from onset to CT, and comorbidities.

| Predictor | OR (95% CI) | p-value |
| --- | --- | --- |
| PVR ($\geq -600$ HU) | 1.131 (1.048–1.221) | 0.002 |
| Age | 1.031 (0.979–1.086) | 0.246 |
| PVR ($\geq -600$ HU) | 1.124 (1.049–1.206) | 0.001 |
| Sex | 0.000 (0.146–1530.796) | 0.252 |
| PVR ($\geq -600$ HU) | 1.126 (1.044–1.214) | 0.002 |
| Number of days from<br>onset to CT | 1.058 (0.837–1.339) | 0.637 |
| PVR ($\geq -600$ HU) | 1.137 (1.045–1.237) | 0.003 |
| Comorbidities (any) | 9.795 (1.432–67.002) | 0.02 |

PVR, pneumonia volume ratio; OR, odds ratio; CT, computed tomography; CI, confidence interval

The evaluation of PVR and CT scores in patients affected by COVID-19-associated pneumonia by Z2 was highly consistent with the visual-evaluation results under the condition of ≥ –600 HU. The AUC and Youden index of the ROC curve by Z2 (≥ –600HU) were 0.881 and 18.69 for PVR, and 0.77 and 7.5 for the CT score, respectively, indicating that they are useful for clinical severity classification.

## Discussion

The chest CT plays a major role in COVID-19 treatment, including severity judgment and prognostic prediction. In clinical practice and in previous studies, the spatial progression of pneumonia on CT has been evaluated with naked eye, and the accuracy and homogeneity have not been ensured.

In this study, we examined the usefulness of determining the severity of COVID-19-associated pneumonia using Z2, an image analysis software widely available in Japan. This methodology can be easily deployed at facilities that have Z2 and thus has high clinical utility.

Several reports evaluated the percentage of lesion area of COVID-19-associated pneumonia in each lobe of the lung visually and scored them to determine the disease severity [6-10]. Yang et al. [6] visually classified the percentage of lesion area in each segment as 0%, <50%, and >50%. Li et al. [7] reported that the percentage of lesion area in each lobe was visually classified as 0%, 0−25%, 25−50%, 50−75%, and 75−100%, and scored on a scale of 0−20. The authors found that the optimal threshold for the severe group was 7.5. Francone et al. [9] used a similar classification, with a mortality risk cut-off of 18. Li et al. [8] also reported scores of 0:0%, 1:<5%, 2:5−25%, 3:25−50%, 4:50−75%, and 5:>75% or higher, with a cut-off score of 7, a sensitivity of 80%, and a specificity of 82.8% for the severely ill group. The cut-off value for clinical severity classification by CT score varies depending on the method and on how the severity is classified.

The CT scores based on visual evaluations that do not require special software or techniques are widely used in clinical settings. This type of evaluation is subjective; however, it has been reported that the inter-evaluator difference is small, and the results of this study are in agreement. However, the score measurement for each lobe in 25–50% increments is troublesome and imposes a burden on the emergency unit staff. Inoue et al. [11] reported that three visual CT score evaluations required 25.7-41.7 s, 27.7-39.5 s, and 48.9-80.0 s, respectively. Novel methods for the quantitative and automated measurement of the spatial progression of COVID-19-associated pneumonia have been reported since the early days of the pandemic [12-21].

Using the commercially available image analysis software, Timaran-Montenegro et al. [12] automatically classified –700 to –1000 HU as normal lung, and –500 to 20 HU as pneumonia regions, and compared the survival vs non-survival groups. The percentage of normal lung was a significant independent factor according to a multinominal logistic analysis. Colombi et al. [13] defined the region of –950 to –700 HU as well as aerated lungs and reported that the measurement by commercially available software and visual measurement were very similar and useful for severity evaluation. In the 10 cases selected in our study, the correlation between the automated measurement by Z2 under the condition of ≥–600 HU and the macroscopic measurement was high: very high for PVR (correlation coefficient 0.842-0.976) and moderate for the CT score (correlation coefficient 0.651-0.696).

As there were no previous reports of using Z2 as a tool to evaluate diseases such as pneumonia with increased lung concentration, the concentration range for pneumonia was determined to be ≥-600 HU in this study, based on the high degree of consistency in terms of visual PVR and CT score.

The range of normal lung, GGO, and consolidation reported in each study using software varied as follows: between: –1000 to –600 HU for normal lung, –750 to –100 HU for GGO, and –399 to –69 HU for consolidation [10-15]. Many previous studies set the lower limit of the GGO range at –800 to –700 HU; however, –600 HU was selected as the lower limit in this study due to the high degree of agreement with the visual findings. This was probably because it is difficult to recognize a faint increase in concentration based on visual evaluation compared to the software-assisted evaluation. It is an advantage of the software-assisted evaluation that it can detect faint concentrations; however, considering that the CT evaluation of COVID-19-associated pneumonia is generally based on visual evaluation, the detection of faint concentrations that are not measurable by visual evaluation leads to clinical discrepancies.

Grassi et al. [14] reported that the percentage of normal lung, emphysema, and consolidation measured by three different software tools were inconsistent. Granata et al. [15] compared the results obtained from two different software tools and reported that the correlation between them was not high enough. The algorithms in which each software is based are different, and therefore comparisons cannot be made under uniform conditions. Z2 is a software tool owned by more than 300 facilities in Japan. Therefore, an assessment method based on the use of Z2 may be immediately available at these facilities and have a high clinical significance. In addition, the introduction of new technologies is time-consuming and expensive.

Okuma et al. [17] reported that the CT score and the percentage of opacity (PVR in this study) obtained using commercially available AI-based software showed a similar AUC; however, in this study the AUC corresponding to PVR and the CT score estimated by Z2 under ≥–600 HU was higher in the case of PVR. Theoretically, the CT scoring method can differ by up to 24% in one lobe at the same point, making it less accurate than PVR. When automated measurement of the same standard becomes widespread, the evaluation by PVR is likely to replace CT scores.

Recently, there have been many reports on the diagnosis and severity assessment of COVID-19-associated pneumonia using AI [16-20]. In a study on COVID-19-associated pneumonia using an AI-based software developed by Ziosoft, the company that developed Z2, Aoki et al. [20] measured the CT lesion extent separately for normal lung, GGO, reticulation, and consolidation. In this study, the pneumonia area was evaluated by combining GGO and consolidation; however, more accurate qualitative and quantitative evaluation will be possible if AI-based software is adopted for this purpose in the future.

In this study, Z2 sometimes misidentified subpleural consolidation as extrapulmonary, requiring manual correction. Inoue et al. [11] reported the measurement errors with the use of U-NET due to the inclusion of atelectasis, fibrosis, and air trapping in the density mask. When a software tool is used, the measurement is carried out automatically; however, the error checking may still need to be performed by human staff.

In this study, we showed the optimal conditions for measuring the PVR and CT score in cases of COVID-19-associated pneumonia using Z2, a widely used image analysis software in Japan, and provided a guideline for clinical severity evaluation based on it. Therefore, defining a Z2-based assessment method has a high clinical significance, and replacing visual evaluation with existing image analysis software represents a way to quickly reduce the burden on clinicians at each facility.

Binomial logistic regression analysis showed no significant effects of age, sex, or time from onset to CT on PVR.

In terms of CT findings, consolidation was significantly higher among the severe group, in agreement with previous reports [9, 19-21]. Several laboratory tests have been reported to be indicators of COVID-19 infection. In our study, both CRP and LDH were significant items, again in agreement with previous reports [22, 23].

The major limitation of this study was the small number of participants at a single facility. The other limitations were that the manual correction of the subpleural consolidation in the Z2 measurement was performed by a single radiologist and the significance of inter-operator differences was not evaluated. Moreover, PVR assumed the area of ≥-600 HU to be a surrogate value for COVID-19 pneumonia, but no histological confirmation was available. The PVR measurements were uniformly performed regardless of the background lesions affecting emphysema, fibrosis, or atelectasis.

In conclusion, we determined the optimal conditions that best approximates visual evaluation for assessing COVID-19-associated pneumonia using Z2, one of the most popular image analysis software tools in Japan and demonstrated that the AUC of PVR was higher than that of CT score in the assessment of clinical severity. The introduction of new technologies is time-consuming and expensive; this method has high clinical utility and can be adopted immediately in any facility where Z2 is available for use.

## Data Availability

Data cannot be disclosed because the subject's consent cannot be obtained.

## Acknowledgments

We would like to thank Dr. Noriko Hida and Dr. Eisuke Inoue for their guidance on the statistical analysis, and Ms. Hokazono and Editage (www.editage.com) for English language editing. We also appreciate the support from our proofreaders and editors. In addition, we are grateful to the clinicians at Fujisawa City Hospital for their insightful advice.

